# Sarcomere Variants of Uncertain Significance identify an Intermediate Clinical Risk Profile in Hypertrophic Cardiomyopathy

**DOI:** 10.64898/2026.03.17.26348662

**Authors:** Hong-Mi Choi, Soo Hyun Seo, In-Chang Hwang, Hyunji Kim, Jee-Soo Lee, Jiesuck Park, Yeonyee E. Yoon, Goo-Yeong Cho, Jaehyun Lim, Soongu Kwak, Jun-Bean Park, Seung-Pyo Lee, Yong-Jin Kim, Moon-Woo Seong, Hyung-Kwan Kim

## Abstract

**Background:** The clinical significance of sarcomere variants of uncertain significance (VUS) in hypertrophic cardiomyopathy (HCM) remains unclear, and VUS are currently regarded as clinically non-actionable despite their increasing prevalence. This study aimed to evaluate genotype–phenotype and genotype–outcome associations according to variant pathogenicity in patients with HCM, with a particular focus on the clinical relevance of sarcomere VUS.

**Methods:** This multicenter retrospective cohort study included 438 patients with HCM who underwent next-generation sequencing-based genetic testing at two tertiary hospitals. Patients were classified into three groups: pathogenic or likely pathogenic (P/LP) variants, VUS, and no sarcomere mutations. Clinical characteristics, imaging phenotypes, and outcomes were compared across groups. The primary endpoint was a composite of cardiovascular death, aborted sudden cardiac death, appropriate implantable cardioverter-defibrillator therapy, and heart transplantation. Time-to-event analyses were performed using Kaplan-Meier methods and Cox proportional hazards models with Firth’s penalized partial likelihood approach.

**Results:** P/LP variants were identified in 171 patients (39.0%) and sarcomere VUS in 159 patients (36.3%). Patients with VUS demonstrated intermediate clinical and phenotypic features between P/LP carriers and genotype-negative patients. Kaplan–Meier analysis showed a graded difference in event-free survival across variant classifications. While VUS were not independently associated with adverse outcomes when modeled as a categorical variable, increasing pathogenicity from genotype-negative to VUS and P/LP variants was associated with a stepwise increase in risk of the primary endpoint (hazard ratio 2.05, 95% confidence interval 1.11–4.16 p=0.019). Identified VUS were preferentially enriched in Z-disc and giant sarcomere scaffolding proteins.

**Conclusion:** Sarcomere VUS represent intermediate characteristics along a continuum of sarcomere dysfunction, associated with distinct phenotypic features and clinical outcomes compared with both P/LP variants and the absence of sarcomere mutations. These findings suggest that sarcomere VUS may not be entirely clinically neutral and should be interpreted within a broader genetic and structural context in patients with HCM.

## INTRODUCTION

Hypertrophic cardiomyopathy (HCM) is one of the most common inherited heart disorders, with an estimated prevalence of approximately 0.2%–0.5% in the general population.^1,2^ Given the associated risk of sudden cardiac death (SCD), particularly in young individuals,^3^ genetic testing and family screening have become integral components of HCM management. Current clinical guidelines recommend genetic testing to facilitate cascade screening of relatives and to support the differential diagnosis of unexplained cardiac hypertrophy.^4,5^

Despite the widespread implementation of genetic testing in HCM, the clinical interpretation of sarcomere variants remains challenging, particularly for variants of uncertain significance (VUS). Pathogenic or likely pathogenic (P/LP) variants are considered causative for HCM, whereas VUS lack sufficient evidence to support pathogenicity and therefore remain clinically non-actionable in current practice.^4,6–8^ However, data from a large multicenter registry suggested that patients carrying sarcomere VUS exhibit an intermediate risk of adverse outcomes between those with pathogenic sarcomere mutations and genotype-negative HCM, raising questions regarding the clinical neutrality of VUS.^9^. In addition, reports indicating that a small portion of sarcomere VUS can be reclassified as P/LP over time further complicate the clinical application of VUS results.^10–12^

Furthermore, the number of reported VUS has increased substantially with the rapid expansion of multigene sequencing panels.^13,14^ This trend has amplified uncertainty in the interpretation of genetic test results and highlights the growing need to better characterize the clinical relevance of VUS in HCM. A more refined assessment of genotype-phenotype and genotype-outcome associations across variant classifications may help bridge the gap between genetic testing and clinical decision-making, particularly for patients carrying VUS.

Accordingly, we sought to determine whether sarcomere VUS represents a phenotypically and prognostically distinct subgroup of HCM rather than a clinically neutral genetic category, by comparing clinical characteristics and outcomes among patients with P/LP variants, VUS, and no sarcomere mutations.

## METHODS

### Study population

This multicenter retrospective cohort study included patients with final diagnosis of HCM who underwent next-generation sequencing (NGS)-based genetic testing for diagnostic evaluation in probands at two tertiary referral hospitals. Cascade testing in relatives was typically performed using targeted sequencing for the familial variant and therefore, was less likely to be captured in this NGS-based cohort.

HCM was diagnosed based on the basis of unexplained left ventricular hypertrophy, defined by maximal end-diastolic wall thickness ≥15mm at any LV segment (or ≥13mm in individuals with a family history of HCM), as assessed by transthoracic echocardiography or cardiac magnetic resonance (CMR), in the absence of other cardiac or systemic conditions capable of causing left ventricular hypertrophy.^4,15,16^ Patients who did not meet diagnostic criteria for HCM on imaging, those diagnosed with HCM phenocopies (including Anderson-Fabry disease, glycogen storage disease, and transthyretin cardiac amyloidosis), and those without available echocardiographic data were excluded. The final study population comprised 438 patients aged ≥15 at the time of genetic testing: 195 patients from Seoul National University Hospital (SNUH) and 243 patients from Seoul National University Bundang Hospital (SNUBH) (**Figure 1**).

**Figure 1.**
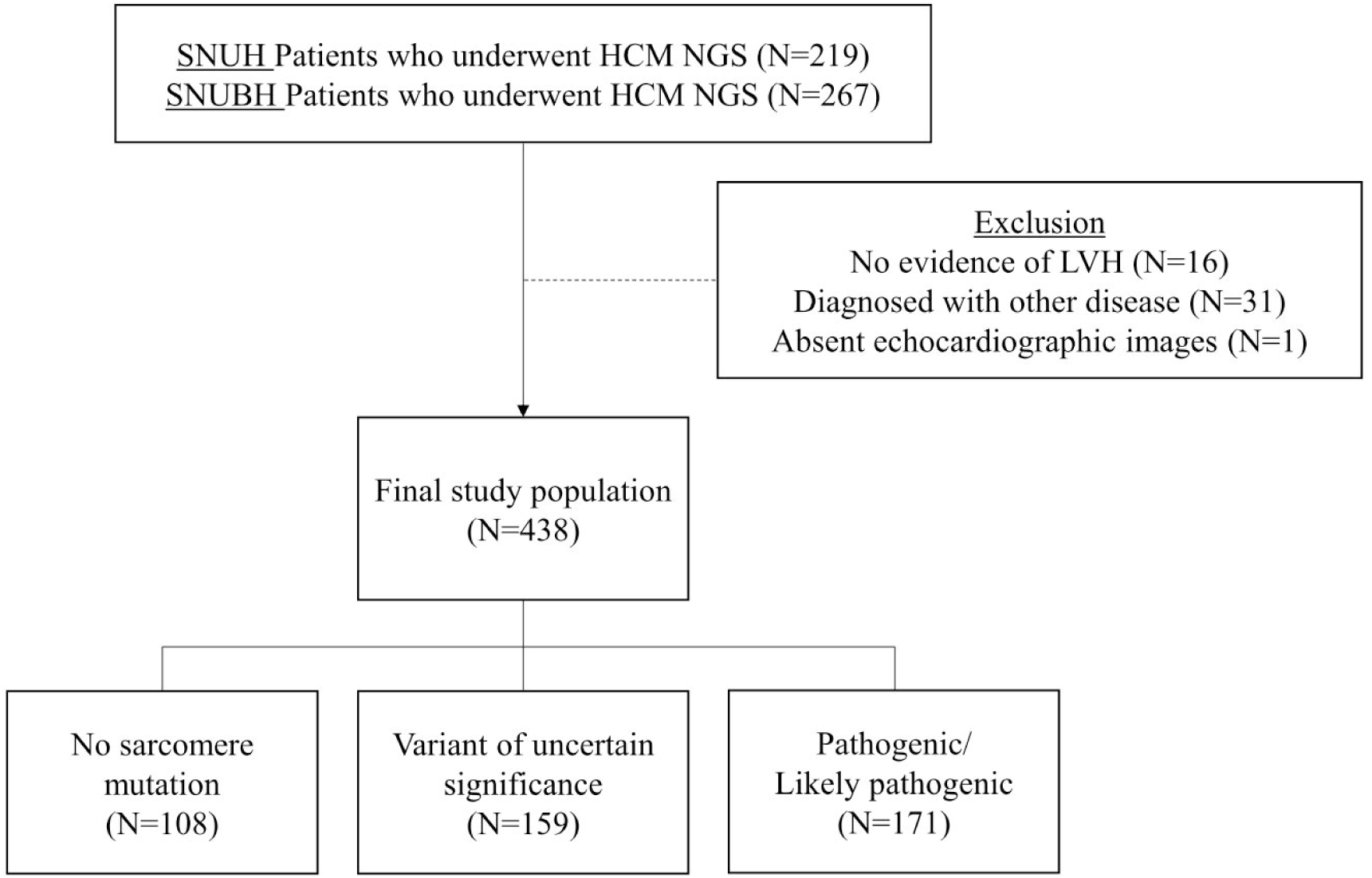
Study population. Abbreviations: HCM, hypertrophic cardiomyopathy, NGS, next generation sequencing, LVH, left ventricular hypertrophy.

### Data collection

Clinical data, including demographic characteristics, established predictors of SCD, diagnostic test results, and outcome-related records, were obtained from electronic health records (EHR). The risk of SCD was estimated using HCM risk-SCD score model, which incorporates age, family history of SCD, maximal left ventricular wall thickness, left atrial diameter, maximal left ventricular outflow tract (LVOT) gradient, non-sustained ventricular tachycardia (VT), and unexplained syncope.^17^ Echocardiographic parameters were collected to assess the phenotypic differences according to genetic status. Echocardiography and CMR images were reviewed in a blinded fashion to clinical and genetic data to classify HCM phenotypes.

The study protocol was approved by the Institutional Review Board of Seoul National University Hospital (IRB No. J-2602-013-1713) and Seoul National University Bundang Hospital (IRB No. B-2004-604-409), with a waiver of written informed consent due to the retrospective study design. All analyses were conducted in accordance with the principles of the Declaration of Helsinki. This observational study follows the STROBE reporting guideline.^18^

### Endpoints

The primary endpoint was an HCM-related major events, which is a composite of cardiovascular (CV) death, aborted SCD, appropriate implantable cardioverter-defibrillator (ICD) discharge, and heart transplantation. SCD-equivalent composite outcome included SCD, aborted SCD, and appropriate ICD discharge. CV death was defined as death resulting from acute myocardial infarction, SCD, heart failure, stroke, CV procedures, CV hemorrhage, and other CV causes.^19^ Aborted SCD was defined as survival following sudden cardiac arrest due to documented ventricular fibrillation or sustained VT requiring external defibrillation or cardiopulmonary resuscitation. ICD therapies delivered for ventricular fibrillation or sustained VT were considered appropriate.^17^ Anti-tachycardia pacing was not considered appropriate ICD therapies.^20^ Clinical events were ascertained from the time of the first HCM diagnosis to the date of the last follow-up or the occurrence of the primary endpoint, whichever came first.

### Gene testing

Genetic testing was performed in patients with a confirmed or suspected diagnosis of HCM, at the discretion of the attending physicians, with patients’ permission to have genetic testing performed. NGS-based panels targeting HCM-associated genes were used. The genes included in the NGS panels are listed in **Supplementary Table S1**. For genotype-phenotype and outcome analyses, variants identified in genes encoding sarcomere structural components were included. Variants in genes associated with HCM phenocopies or non-sarcomere cardiomyopathies were excluded. Sarcomere genes were further classified, according to their predominant structural roles, into thick filament (A-band), thin filament (I-band), Z-disc, and giant sarcomere scaffolding protein groups, as summarized in **Supplementary Table S2**.

The pathogenicity of detected variants was evaluated according to the 2015 American College of Medical Genetics and Genomics and the Association for Molecular Pathology (ACMG/AMP) guidelines with application of the ClinGen Cardiomyopathy Expert Panel gene-specific specifications.^21,22^ Variant interpretation included assessment of gene-specific evidence supporting HCM causality, including population allele frequencies, in silico predictive data, functional studies, and available segregation data. Based on this evaluation, variants were classified as P/LP, VUS, or benign/likely benign by laboratory medicine physicians. In this study, the VUS group was defined as individuals carrying one or more VUS in the absence of any P/LP variants. VUS burden was defined as the cumulative number of sarcomere gene VUS per individual and was categorized as 0, 1, 2, or ≥3 variants.

### Statistical analysis

Continuous variables are presented as means with standard deviations or medians with interquartile ranges, as appropriate, and categorical variables as frequencies with percentages. Baseline characteristics were compared across sarcomere variant classification using one-way analysis of variance or the Kruskal-Wallis test for continuous variables and the chi-square test or Fisher’s exact test for categorical variables, as appropriate.

Time-to-event outcomes were analyzed using Kaplan–Meier survival curves, with between-group differences assessed using the log-rank test. Cox proportional hazards regression models with Firth’s penalized partial likelihood approach were used to estimate hazard ratios (HRs) and profile-likelihood–based 95% confidence intervals (CIs), given the limited number of outcome events and the potential for small-sample bias.^23^ Multivariable Cox models were adjusted for age and sex and stratified by institution. Morphological subtype, maximal wall thickness, LVOT obstruction, SCD-related factors, and comorbidities were not included in the adjusted models because these variables were considered potential downstream manifestations of genotype^10,24^. The proportional hazards assumption was assessed using Schoenfeld residuals and was not violated.

Sensitivity analyses were performed restricting the analysis to sarcomere genes with strong evidence for pathogenicity (i.e., *MYH7*, *MYBPC3*, *TNNI3*, *TNNT2*, *TPM1*, *MYL2*, *MYL3*, and *ACTC1)*, which are recognized to explain approximately 90% of genotype-positive HCM cases.^4,5^ Statistical significance was defined using a two-sided p-value <0.05.

All analyses were performed using R, version 4.4.1 (https://www.R-project.org).

## RESULTS

### Baseline characteristics

Baseline clinical characteristics of 438 patients are summarized in **Table 1**. The mean age at the time of genetic testing was 56.3 ± 14.2 years, and 68.0% of the patients were male. A family history of HCM was present in 24 patients (5.5%), and 60 patients (13.7%) had family history of SCD. Septal HCM was the most common morphological subtype (44.0%), followed by apical HCM (22.4%). The mean maximal left ventricular wall thickness was 19.1 ± 4.2 mm, and LVOT obstruction was present in 90 patients (20.5%). The mean left ventricular ejection fraction was 64.0 ± 7.2%. Left atrial enlargement was common, with a mean left atrial diameter of 43.2 ± 7.6 mm and a left atrial volume index of 46.6 ± 19.6 mL/m^2^. Normal diastolic function was observed in 25.8% of patients.

**Table 1.** Baseline characteristics.

|  | N (%) or Mean±SD |
| --- | --- |
| Age at the time of genetic testing | 56.3±14.2 |
| Male sex | 298 (68.0%) |
| Morphological subtype of HCM |  |
| Septal | 196 (44.7%) |
| Apical | 98 (22.4%) |
| Diffuse | 61 (13.9%) |
| Mixed | 68 (15.5%) |
| Others/atypical | 15 (3.4%) |
| Family history of HCM | 24 (5.5%) |
| *SCD-related factors* |  |
| Family history of SCD | 60 (13.7%) |
| Nonsustained VT | 111 (25.3%) |
| Syncope | 43 (9.8%) |
| HCM risk-SCD score | 2.9±2.4 |
| ICD implantation | 71 (16.2%) |
| *Comorbidity* |  |
| Diabetes mellitus | 52 (11.9%) |
| Hypertension | 176 (40.2%) |
| Chronic kidney disease | 10 (2.3%) |
| End stage kidney disease | 2 (0.5%) |
| Atrial fibrillation | 38 (8.7%) |
| Prior history of PCI | 2 (0.5%) |
| Prior history of stroke | 28 (6.4%) |
| *Genetic testing (limited to sarcomere genes)* |  |
| Pathogenic/Likely Pathogenic variants | 171 (39.0%) |
| Variants of uncertain significance | 159 (36.3%) |
| No variants | 108 (24.7%) |
| *Echocardiography* |  |
| Maximal LV wall thickness (mm) | 19.1±4.2 |
| Maximal LVOT pressure gradient (mmHg) | 22.5±37.8 |
| LVOT obstruction | 90 (20.5%) |
| LV end-diastolic diameter (mm) | 45.6±5.4 |
| LV end-systolic diameter (mm) | 27.8±5.0 |
| LV ejection fraction (%) | 64.0±7.2 |
| E (m/s) | 0.6±0.2 |
| A (m/s) | 0.6±0.2 |
| E/e' | 13.7±5.7 |
| LV GLS (absolute value) | 14.1±4.6 |
| LA diameter (mm) | 43.2±7.6 |
| LAVI (ml/m<sup>2</sup>) | 46.6±19.6 |
| Diastolic function |  |
| Normal diastolic function | 113 (25.8%) |
| Grade 1 (abnormal relaxation) | 202 (46.1%) |
| Grade 2 (pseudonormal pattern) | 99 (22.6%) |
| Grade 3 (restrictive filling pattern) | 14 (3.2%) |
| Absence of A wave | 10 (2.3%) |
| PASP (mmHg) | 31.6±7.5 |

Abbreviations: GLS, global longitudinal strain; LA, left atrial; LAVI, left atrial volume index; LV, left ventricular; LVOT, left ventricular outflow tract; PASP, pulmonary arterial systolic pressure; PCI, percutaneous coronary intervention; SCD, sudden cardiac death; SD, standard deviation; VT, ventricular tachycardia. LVOT obstruction refers to maximal LVOT pressure gradient $\geq 30$ mmHg.

P/LP sarcomere gene variants were identified in 171 patients (39.0%), whereas VUS in the absence of P/LP variants were identified in 159 patients (36.3%). Clinically insignificant sarcomere gene variants (benign or likely benign) or no sarcomere mutations were observed in 108 patients (24.7%). Among P/LP carriers, variants were most frequently identified in *MYBPC3* (85 patients, 48.9%), followed by *MYH7* (49 patients, 28.2%) (**Supplementary Table S3**). Three patients carried two P/LP variants in different sarcomere genes, and two patients carried two distinct P/LP variants in *MYBPC3*. The most frequently identified P/LP variant was *TNNI3* c.434G>A (p.Arg145Gln), which was present in 19 patients. No P/LP variants or VUS were identified in *DES* in this cohort.

### Baseline characteristics by sarcomere variant classification

Baseline characteristics differed significantly according to sarcomere variant classification (**Table 2**). Patients with P/LP variants were younger at the time of genetic testing compared with those carrying VUS or no mutations (52.2 ± 14.4, 58.3 ± 13.8, and 60.0 ± 12.8 years, respectively, p<0.001) and were less likely to be male. The distribution of sarcomere genetic status was significantly different across the institution. Phenotypes characterized by septal hypertrophy, including septal and mixed forms, predominated among patients with P/LP sarcomere variants, accounting for approximately 74% of cases. In contrast, apical or diffuse HCM was more frequently observed in patients with VUS or without sarcomere gene mutations.

**Table 2.** Baseline characteristics stratified by sarcomere variant classification.

|  | No sarcomere<br>mutation<br>(N=108) | Sarcomere<br>VUS<br>(N=159) | Sarcomere<br>P/LP<br>(N=171) | p |
| --- | --- | --- | --- | --- |
| Age at the time of genetic testing | 60.0±12.8 | 58.3±13.8 | 52.2±14.4 | <0.001 |
| Male sex | 81 (75.0%) | 116 (73.0%) | 101 (59.1%) | 0.005 |
| Institution |  |  |  | <0.001 |
| SNUH | 83 (76.9%) | 72 (45.3%) | 88 (51.5%) |  |
| SNUBH | 25 (23.1%) | 87 (54.7%) | 83 (48.5%) |  |
| Morphological subtype of HCM |  |  |  | <0.001 |
| Septal | 35 (32.4%) | 71 (44.7%) | 90 (52.6%) |  |
| Apical | 35 (32.4%) | 41 (25.8%) | 22 (12.9%) |  |
| Diffuse | 21 (19.4%) | 23 (14.5%) | 17 (9.9%) |  |
| Mixed | 16 (14.8%) | 16 (10.1%) | 36 (21.1%) |  |
| Others/atypical | 1 (0.9%) | 8 (5.0%) | 6 (3.5%) |  |
| Family history of HCM | 2 (1.9%) | 5 (3.1%) | 17 (9.9%) | 0.004 |
| *SCD-related factors* |  |  |  |  |
| Family history of SCD | 16 (14.8%) | 16 (10.1%) | 28 (16.4%) | 0.231 |
| Nonsustained VT | 15 (13.9%) | 36 (22.6%) | 60 (35.1%) | <0.001 |
| Syncope | 9 (8.3%) | 17 (10.7%) | 17 (9.9%) | 0.815 |
| HCM risk-SCD score | 2.2± 1.2 | 2.7± 2.0 | 3.6± 3.1 | <0.001 |
| ICD implantation | 11 (10.2%) | 16 (10.1%) | 44 (25.7%) | <0.001 |
| *Comorbidities* |  |  |  |  |
| Diabetes mellitus | 18 (16.7%) | 19 (11.9%) | 15 (8.8%) | 0.139 |
| Hypertension | 56 (51.9%) | 65 (40.9%) | 55 (32.2%) | 0.005 |
| Chronic kidney disease | 3 (2.8%) | 4 (2.5%) | 3 (1.8%) | 0.749 |
| End stage kidney disease | 0 (0.0%) | 1 (0.6%) | 1 (0.6%) | 0.719 |
| Atrial fibrillation | 7 (6.5%) | 16 (10.1%) | 15 (8.8%) | 0.593 |
| Prior history of PCI | 2 (1.9%) | 0 (0.0%) | 0 (0.0%) | 0.046 |
| Prior history of stroke | 15 (13.9%) | 7 (4.4%) | 6 (3.5%) | 0.001 |
| *Echocardiographic parameters* |  |  |  |  |
| Maximal LV wall thickness (mm) | 18.3± 3.3 | 18.5± 4.0 | 20.2± 4.6 | <0.001 |
| Maximal LVOT pressure gradient (mmHg) | 22.8±38.7 | 23.0±42.7 | 21.8±32.1 | 0.957 |
| LVOT obstruction | 23 (21.3%) | 27 (17.0%) | 40 (23.4%) | 0.346 |
| LV end-diastolic diameter (mm) | 46.5± 5.1 | 46.6± 5.3 | 44.1± 5.5 | <0.001 |
| LV end-systolic diameter (mm) | 28.6± 4.8 | 28.4± 4.6 | 26.8± 5.3 | 0.003 |
| LV ejection fraction (%) | 64.9± 6.5 | 64.0± 6.3 | 63.5± 8.2 | 0.331 |
| E (m/s) | 0.7± 0.2 | 0.6± 0.2 | 0.6± 0.2 | 0.899 |
| A (m/s) | 0.7± 0.2 | 0.6± 0.2 | 0.6± 0.2 | <0.001 |
| E/e' | 13.2± 5.3 | 13.5± 5.6 | 14.1± 5.9 | 0.432 |
| LV GLS (absolute value) | 13.7± 4.1 | 14.2± 4.5 | 14.1± 5.0 | 0.830 |
| LA diameter (mm) | 42.2± 6.5 | 43.9± 8.1 | 43.1± 7.8 | 0.220 |
| LAVI (ml/m<sup>2</sup>) | 43.5±13.9 | 46.8±19.8 | 48.5±22.3 | 0.132 |
| Diastolic function |  |  |  | 0.089 |
| Normal diastolic function | 23 (21.3%) | 43 (27.0%) | 47 (27.5%) |  |
| Grade 1 | 64 (59.3%) | 71 (44.7%) | 67 (39.2%) |  |
| Grade 2 | 16 (14.8%) | 37 (23.3%) | 46 (26.9%) |  |
| Grade 3 | 2 (1.9%) | 4 (2.5%) | 8 (4.7%) |  |
| Absence of A wave | 3 (2.8%) | 4 (2.5%) | 3 (1.8%) |  |
| Diastolic dysfunction ≥2 | 18 (16.7%) | 41 (25.8%) | 54 (31.6%) | 0.021 |
| PASP (mmHg) | 30.3± 7.4 | 31.8± 7.7 | 32.2± 7.4 | 0.228 |

Abbreviations: P, pathogenic variants; LP, likely pathogenic variants; VUS, variants of uncertain significance. Other abbreviations are defined in the footnote of Table 1.

Patients with P/LP variants had a higher prevalence of nonsustained VT and higher HCM-risk SCD scores compared with those without sarcomere mutations, which was associated with a higher rate of ICD implantation. In addition, patients carrying P/LP variants showed greater maximal left ventricular wall thickness and smaller left ventricular dimensions than those with VUS and without sarcomere mutation. Diastolic dysfunction showed a graded pattern across variant classifications, with intermediate values observed in patients with VUS. **Figure 2** illustrates phenotypic heterogeneity according to sarcomere variant classification, highlighting the intermediate clinical profiles of patients harboring VUS.

**Figure 2.**
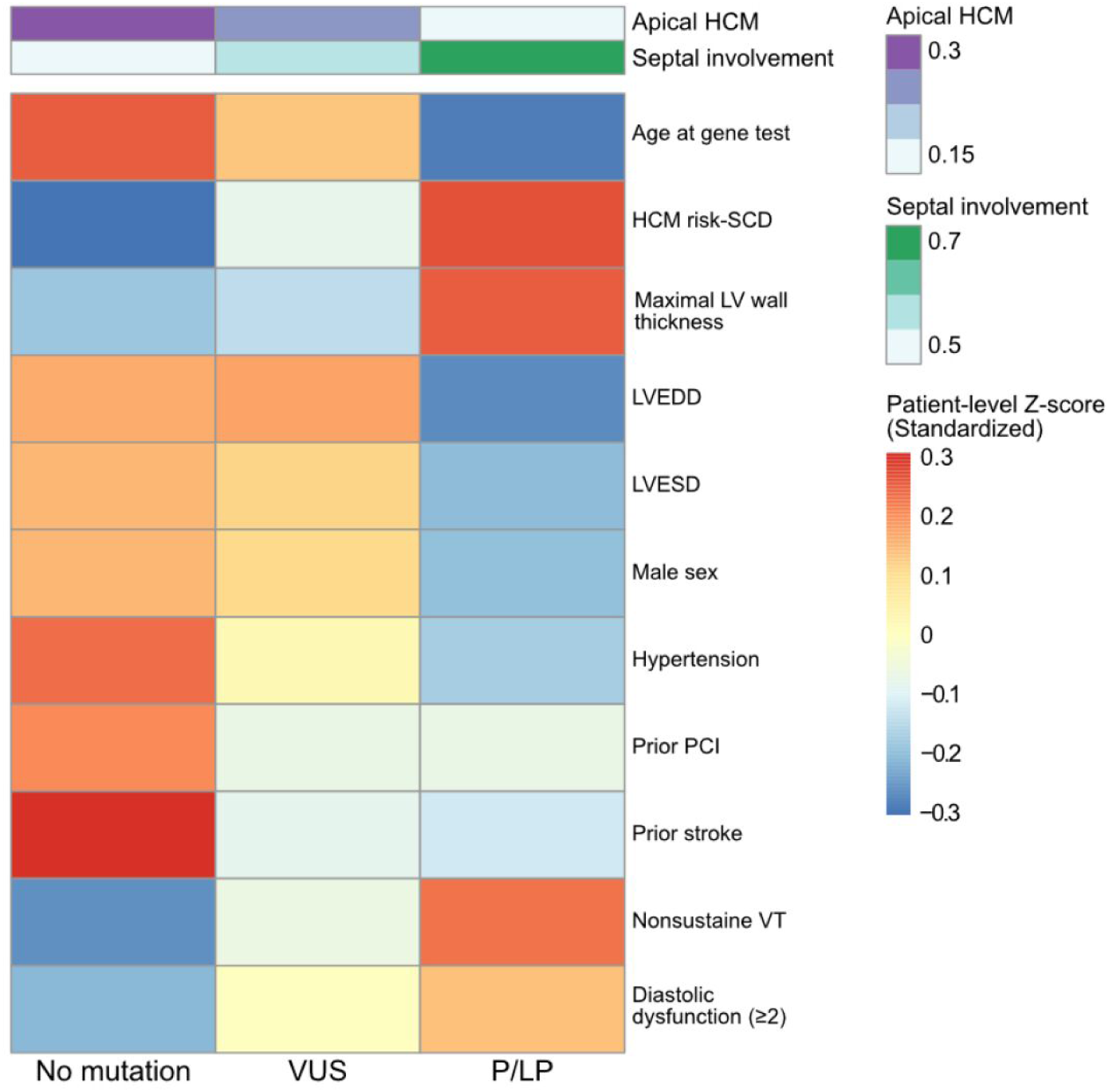
Standardized heatmap of clinical and echocardiographic features according to sarcomere genetic status. Column annotations indicate the proportion of apical and septal-involving phenotypes within each group. For each variable, patient-level Z-scores were first calculated by standardizing the raw values across the entire cohort. Each cell in the heatmap then represents the mean of these patient-level Z-scores within the respective genetic group. Positive values indicate higher levels or prevalence relative to the overall cohort average, whereas negative values indicate lower levels. Binary variables were standardized in the same manner. Abbreviations: HCM, hypertrophic cardiomyopathy, SCD, sudden cardiac death, LV, left ventricular, LVEDD, left ventricular end-diastolic dimension, LVESD, left ventricular end-systolic dimension, PCI, percutaneous coronary intervention, VT, ventricular tachycardia, VUS, variants of uncertain significance, P/LP, pathogenic/likely pathogenic.

### Clinical outcomes according to sarcomere genetic status

Primary endpoint occurred in 29 patients (6.6%), including 20 events occurred among carriers of P/LP sarcomere variants. The SCD-equivalent composite outcome developed in 24 patients (5.5%), of whom 16 were P/LP variants carriers. The median follow-up duration was 4.9 [1.5–8.7] years in patients without sarcomere mutations, 6.8 [3.2–12.1] years in those with sarcomere VUS, and 6.0 [2.6–10.0] years in those with P/LP variants. The incidence of the primary endpoint and SCD-equivalent composite outcome was significantly higher in patients with P/LP sarcomere variants compared with those without sarcomere mutations (*p* = 0.001 and 0.008, respectively, **Table 3**).

**Table 3.**
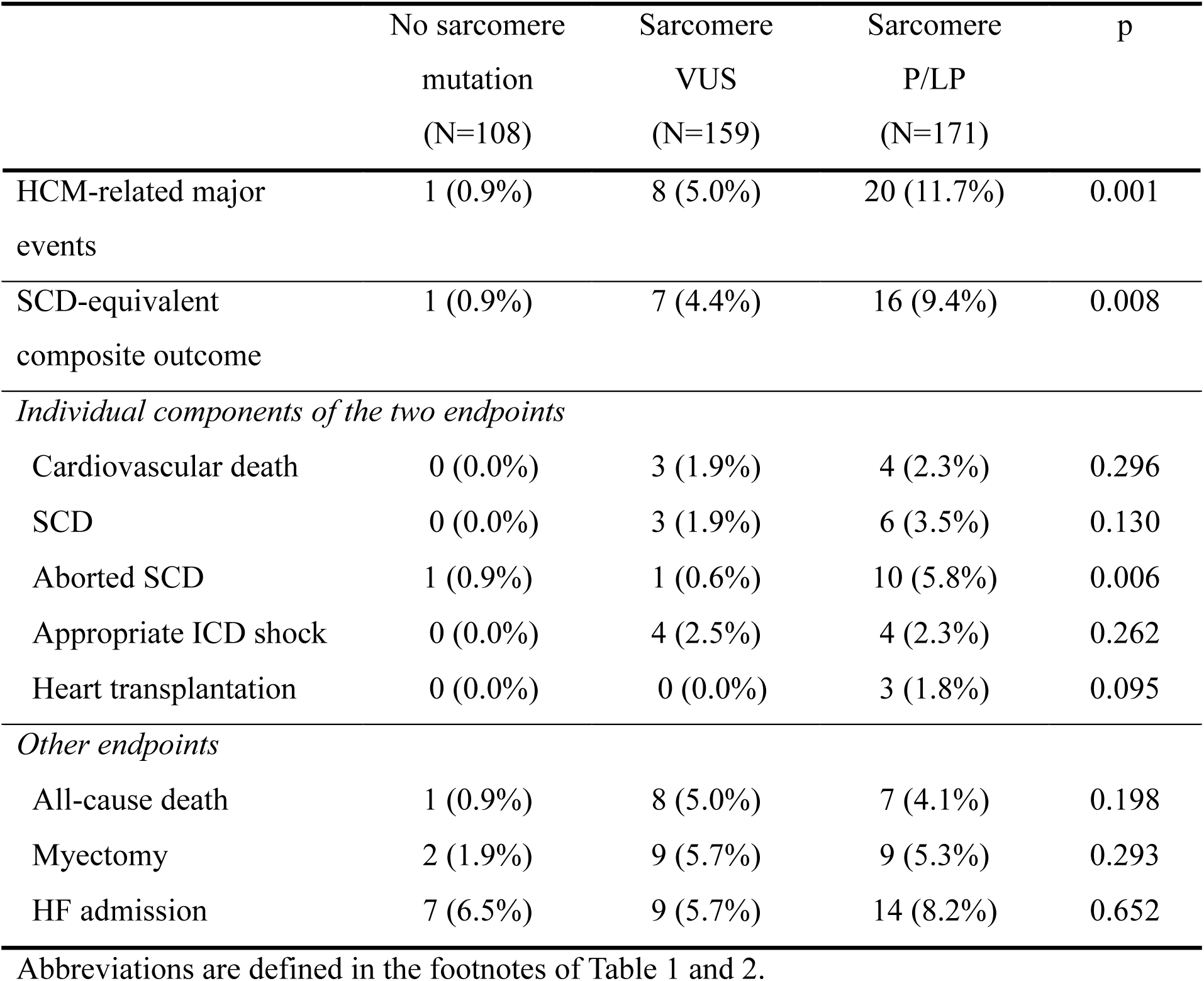
Outcomes stratified by sarcomere variant classification.

Kaplan–Meier analysis showed a significant difference in event-free survival across sarcomere variant classifications for the primary endpoint (log-rank *p* = 0.0026). For the SCD-equivalent endpoint, a similar graded pattern was observed, although the difference did not reach statistical significance (log-rank *p* = 0.083) (**Figure 3**). Among the individual components of these two endpoints, aborted SCD occurred more frequently in patients with P/LP sarcomere gene variants, whereas no statistically significant differences were observed for the other components (**Table 3**).

**Figure 3.**
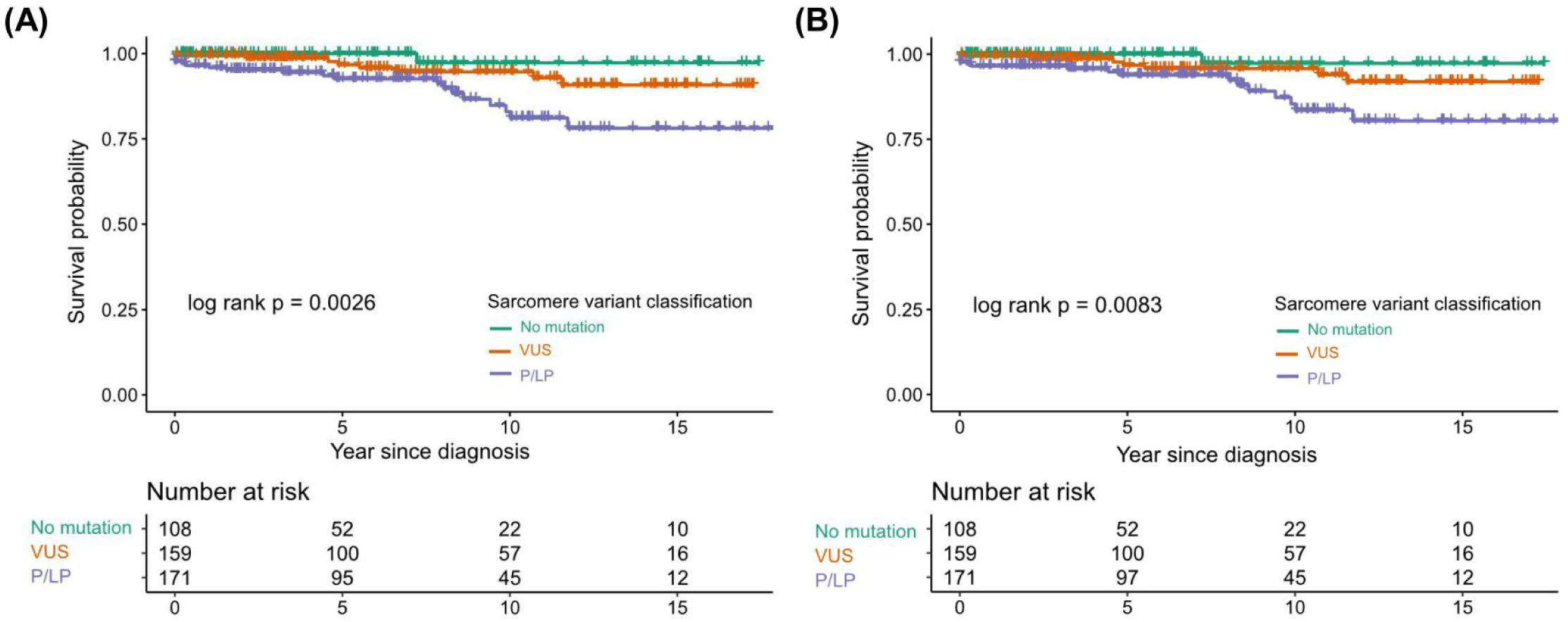
Kaplan–Meier curves according to sarcomere genetic status for event-free survival from (A) HCM-related major events and (B) SCD-equivalent composite outcome. HCM-related major events is a composite of cardiovascular death, aborted sudden cardiac death, appropriate implantable cardioverter-defibrillator shock, and heart transplantation. SCD-equivalent composite outcome includes SCD, aborted SCD, and appropriate ICD discharge. Differences between groups were assessed using the log-rank test. VUS, variant of uncertain significance, P/LP, pathogenic/likely pathogenic variant.

To further quantify the association between sarcomere genetic status and clinical outcomes, multivariable Firth-penalized Cox proportional hazards models were constructed. After adjustment for age and sex and stratification by institution, P/LP variants were independently associated with an increased risk of the primary endpoint when modelled as separate variables compared with no sarcomere mutation (HR 4.57, 95% CI 1.11–42.02, p=0.034), whereas sarcomere VUS was not (**Table 4**). When sarcomere variant classification was modelled as an ordinal variable reflecting increasing pathogenic potential (no sarcomere mutation → VUS → P/LP variants), a significant dose-response relationship was observed, with progressively higher risk of the primary endpoint across increasing levels of genetic burden (HR 2.05, 95% CI 1.11–4.16, p=0.019, **Table 4**).

**Table 4.**
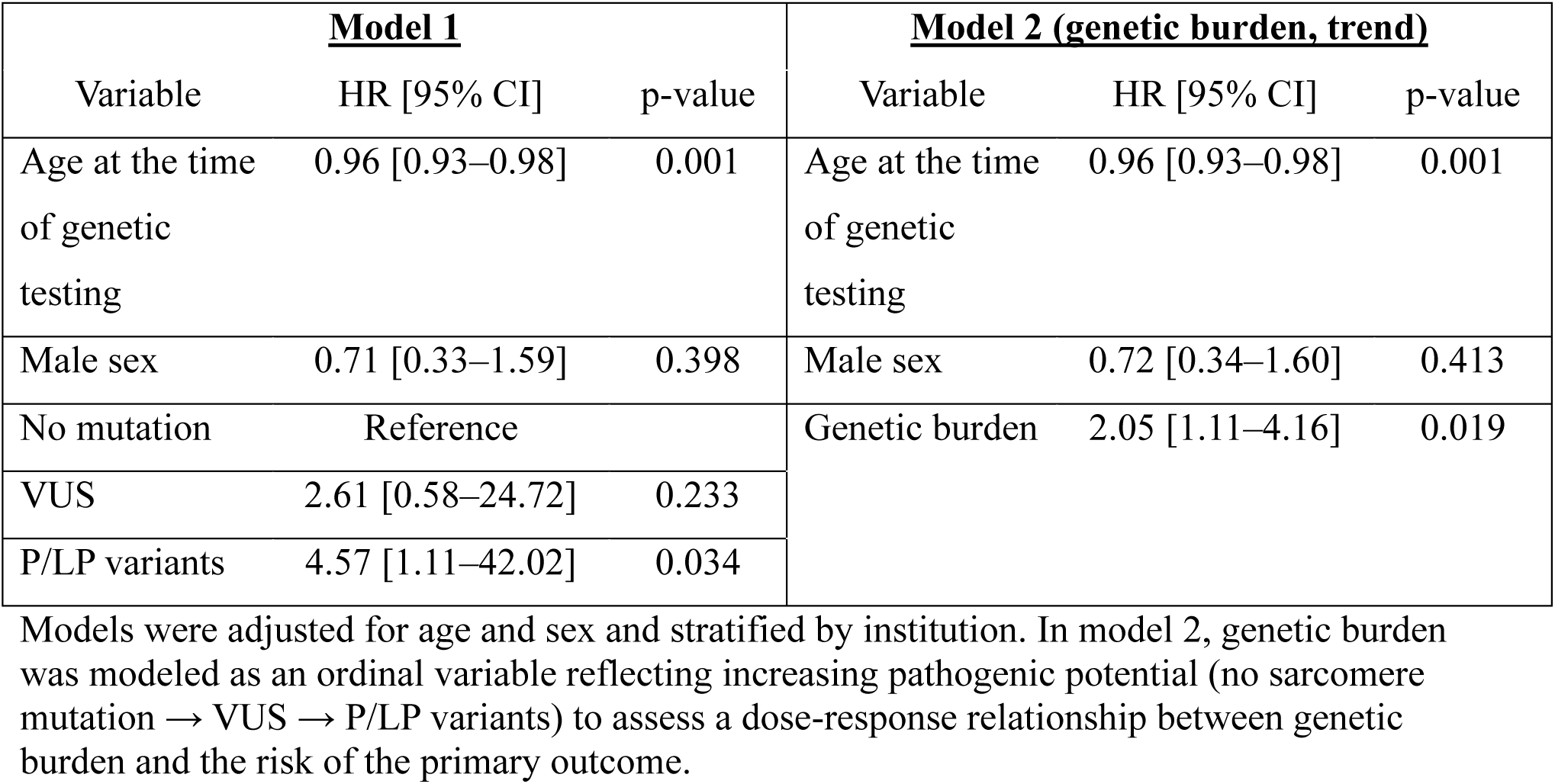
Multivariable Firth-penalized Cox regression analyses for HCM-related major events.

### Association of VUS burden with clinical outcomes

To further evaluate the clinical relevance of VUS, subgroup analyses were performed according to VUS burden. Among patients carrying only sarcomere gene VUS, the proportion of the primary endpoint increased with higher VUS burden, occurring in 4.1%, 5.0%, and 9.1% of patients with 1, 2, and ≥3 VUS, respectively. However, these differences did not reach statistical significance (Fisher’s exact test p = 0.51). Among sarcomere P/LP carriers, the presence of additional VUS was not associated with a higher risk of the primary endpoint (Fisher’s exact test p = 0.47, **Supplementary Table S4**).

### Distribution of sarcomere gene variants by structural category

When sarcomere variants were stratified by sarcomere structural category, substantial differences in pathogenicity classification of sarcomere variant were observed (**Figure 4**). Variants affecting thick and thin filament proteins were predominantly classified into P/LP, whereas nearly all variants in giant scaffolding proteins were classified as VUS. No P/LP variants were identified in Z-disc proteins.

**Figure 4.**
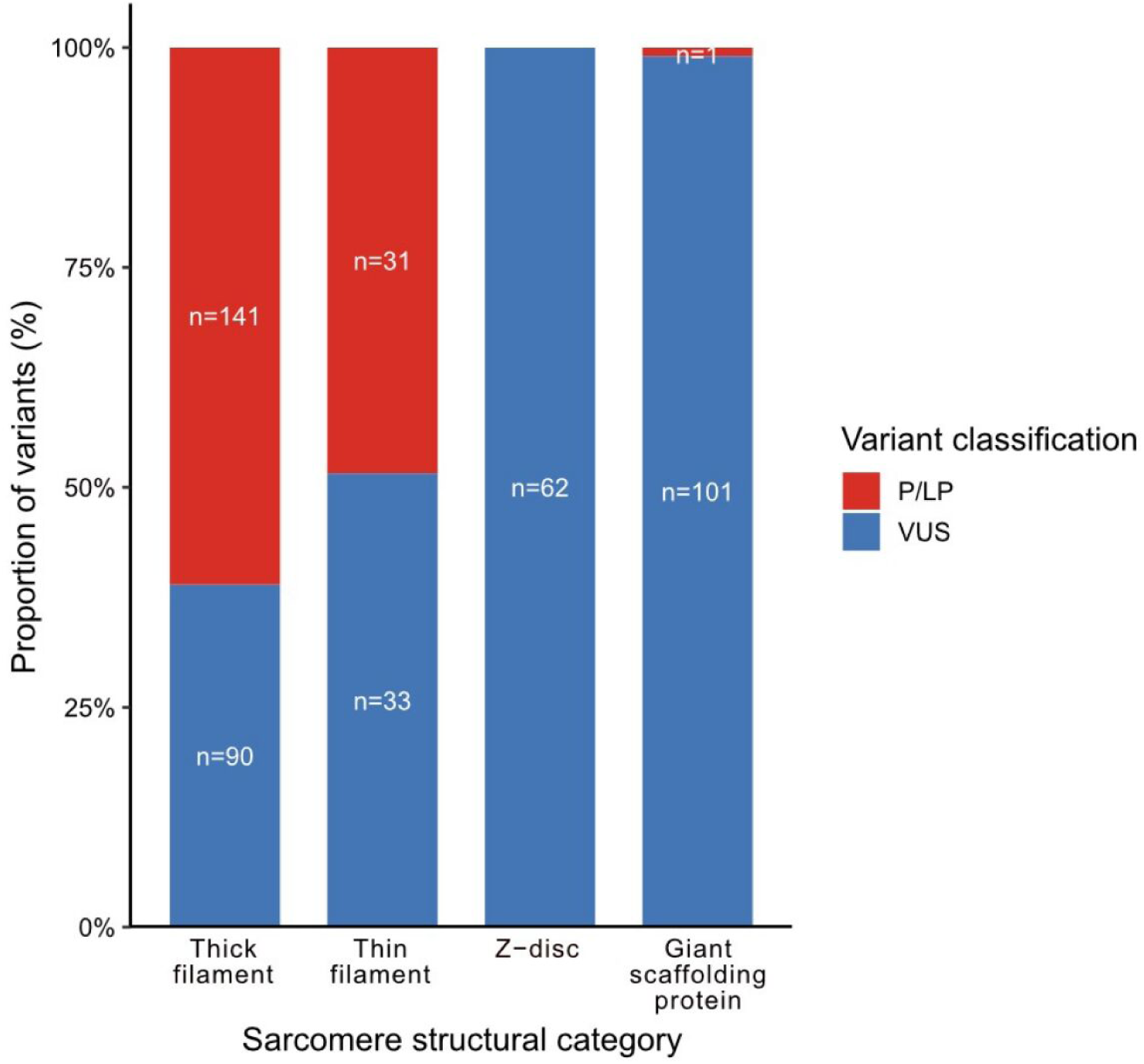
Distribution of sarcomere gene variants by structural category. Stacked bars represent the proportion of pathogenic/likely pathogenic (P/LP) variants and variants of uncertain significance (VUS) across sarcomere structural categories. Absolute numbers of variants are shown within each bar. VUS were disproportionately enriched in Z-disc and giant scaffolding proteins compared with thick and thin filament genes.

### Sensitivity analysis

In the sensitivity analysis restricted to genes with strong evidence for HCM, P/LP variants were identified in 168 patients (38.4%), whereas VUS in the absence of P/LP variants were identified in 77 patients (17.6%). The findings were consistent with those of the primary analysis, showing slightly more prominent pathogenicity of P/LP variants and a trend toward greater phenotypic similarity between patients with VUS and those with P/LP variants, as evidenced by genotype-phenotype and genotype-outcome associations **(Supplementary Table S5-S6 and Supplementary Figure S1-S2**).

## DISCUSSION

In this multicenter cohort of patients with HCM, we observed distinct differences in baseline characteristics, cardiac phenotype, and clinical outcomes according to sarcomere variant classification. Patients carrying P/LP variants exhibited the highest risk profile, whereas carriers of VUS demonstrated intermediate phenotypic and clinical features between P/LP carriers and patients without sarcomere mutations. Notably, among patients carrying only sarcomere VUS, a graded increase in the occurrence of the primary endpoint was observed with increasing VUS burden, suggesting a potential cumulative effect of multiple VUS. In addition, by including sarcomere variants beyond thick and thin filament proteins, this study demonstrated a marked enrichment of VUS in Z-disc and giant scaffolding proteins. These findings indicate that sarcomere VUS do not represent a homogeneous or clinically neutral category and underscore the need for a more nuanced interpretation of variant classification in patients with HCM.

The clinical implications of VUS have been increasingly investigated in patients with HCM. Ho *et al*. well demonstrated that patients carrying sarcomere VUS exhibit an intermediate phenotype, characterized by earlier disease onset and worse clinical outcomes compared with genotype-negative patients.^9^ Findings from this large-scale registry with long-term follow-up suggest that a subset of VUS may confer pathogenic potential despite not meeting the stringent criteria of ACMG guidelines. However, the reproducibility of these observations across independent cohorts and the mechanisms by which VUS contribute to phenotype and pathogenic heterogeneity in HCM remain incompletely understood. Moreover, as the NGS panel expands, increasing VUS rates make VUS interpretation more complicated.

In the present study, sarcomere genetic status demonstrated a graded association with clinical outcomes and disease expression. Patients carrying VUS exhibited clinical characteristics that were intermediate between genotype-negative individuals and P/LP variant carriers. Specifically, the distribution of morphological subtypes and comorbidities were significantly different across the variant classification, and the HCM risk-SCD score in patients with VUS was located between those of P/LP carriers and genotype-negative patients, indicating that VUS status aligns with established SCD risk stratification frameworks. For a prognostic perspective, while only P/LP variants were significantly associated with increased risk compared with no sarcomere mutation in multivariable Firth-penalized Cox models, modelling variant classification as an ordinal variable revealed a significant dose-response relationship. Similar trend was observed in the sensitivity analysis, emphasizing the robustness of our findings.

Hernandez *et al*. recently introduced the concept of intermediate-effect variants (IEVs) to describe variants with moderate effect sizes that modulate disease expression.^25^ Our findings align with this framework, suggesting that the VUS identified in our cohort, particularly those enriched in Z-disc and giant scaffolding proteins including FLNC, TTN, and OBSCN, function clinically as IEVs. Unlike the binary classification of pathogenic or benign, these variants confer a quantitative risk that lies between genotype-negative and P/LP positive cases, warranting a shift towards a continuous risk model in HCM management.

Despite these associations, patients carrying VUS follow distinct management pathways compared with P/LP carriers. Contemporary HCM guidelines do not recommend cascade genetic testing for family members of patients with VUS.^4^ In addition, recent guidelines for SCD prevention consider ICD implantation reasonable in patients with intermediate 5-year risk score of SCD and P/LP sarcomere mutation (Class IIA), but not in those carrying VUS.^8^ Because sarcomere VUS are regarded as equivalent to genotype-negative status, patients with VUS may have limited access to emerging precision medicine strategies, including gene-based therapy.^26,27^

Reclassification of VUS may be achieved through multiple approaches, including computational modeling, expanding population- and disease-specific databases, segregation analyses, functional assays, and RNA studies.^13,28^ However, the systematic application of these approaches remain challenging in real-world practice and requires additional effort and cost.^29^ Moreover, the number of VUS is expected to increase as the sequencing technologies becomes faster and more affordable.^30^ Consequently, newly identified variants in additional sarcomere genes are more likely to be categorized as VUS. In this context, our findings underscore the growing clinical implications of VUS in HCM.

Several biological and structural mechanisms may underlie the intermediate clinical phenotype observed in patients carrying sarcomere VUS. First, VUS are more likely to represent missense or other non-truncating variants with modest functional effects, which may be insufficient to cause overt disease independently but can contribute to disease expression in a context-dependent manner ^19,31^ Second, VUS were disproportionately enriched in genes encoding Z-disc and giant scaffolding proteins in this study. Current guideline recognize only eight sarcomere genes with strong evidence for disease causality (i.e., *MYH7*, *MYBPC3*, *TNNI3*, *TNNT2*, *TPM1*, *MYL2*, *MYL3*, and *ACTC1*),^4,32^ whereas other sarcomere genes were are less frequently included in the multigene panels. As a result, variants identified in these genes are less likely to accumulate sufficient evidence for P/LP classification. Furthermore, because Z-disc and giant scaffolding proteins primarily play regulatory roles rather than directly generating contractile force, variants in these domains may lead to subtle alterations in sarcomere organization, force transmission, or intracellular signaling, resulting in incomplete penetrance and variable expressivity.^33–36^ Third, limitations in current variant classification system may contribute to the high prevalence of VUS, as pathogenicity assignment relies heavily on segregation data and functional assays, which are less well established for non-thick filament sarcomere proteins.^37^

From a clinical perspective, sarcomere VUS should be interpreted within a broader biological and structural context rather than being regarded as uniformly benign. At the same time, caution is warranted to avoid overinterpretation of VUS, which could lead to unnecessary testing and wasting of healthcare resources. In parallel, continued efforts to improve VUS interpretation are essential, through advances in variant reclassification methodologies and large-scale biobank initiatives linking genomic and phenotypic data. Improved understanding of the relationship between sarcomere VUS and clinical phenotype may ultimately help bridge the gap between genetic uncertainty and clinical decision-making in patients with HCM.

This study has several limitations. First, genetic testing was not performed in all patients with HCM, resulting in a relatively limited sample size for genotype-phenotype and outcome analyses. Accordingly, selection bias cannot be completely excluded, and the distribution of sarcomere variant classifications in this cohort may not fully represent that of the overall HCM population. Specifically, the number of HCM-related major events among genotype-negative patients was very small, which may have contributed to imprecision in risk estimates and wide confidence intervals. The observed event rate in this group was lower than that reported in prior cohorts,^9^ potentially reflecting selection bias owing to the shorter follow-up period. Although we introduced Firth’s penalization approach to survival analysis to mitigate bias arising from a small number of events, these findings should be interpreted with caution and warrant validation in larger, more diverse populations with long-term follow-up. Second, the composition of NGS panels varied over time and across institutions, which might have influenced the observed distribution of sarcomere variants. Although all panels consistently included eight sarcomere genes with strong evidence for HCM causality, caution is warranted when generalizing the findings due to non-standardized panel composition. Third, since proband status and family relatedness were not systematically captured, a small proportion of relatives may have been included. Lastly, despite a median follow-up duration of 6.1 [interquartile range, 2.5–10.5] years, this period may be insufficient to capture the full spectrum of long-term and progressive clinical outcomes in HCM. To further clarify the clinical significance of sarcomere VUS, large-scale prospective studies using standardized gene panels or whole-genome sequencing with longer follow-up are warranted.

In conclusion, sarcomere VUS represent an intermediate category along a continuum of sarcomere dysfunction, characterized by distinct phenotypic features and clinical outcomes that differ from both P/LP variants and the absence of sarcomere mutations. The observed trend toward a higher incidence of adverse outcomes with increasing VUS burden, together with the preferential localization of VUS in Z-disc and giant sarcomere scaffolding proteins, underscores the potential biological relevance of these variants. These findings highlight that sarcomere VUS may have clinical relevance beyond a strictly neutral classification and warrant cautious interpretation within a broader genetic and structural context in patients with HCM.

## Data Availability

The datasets analyzed during the current study are not publicly available due to institutional restrictions. A minimally anonymized dataset can be requested by contacting the corresponding authors.

## Author Contributions

All authors were involved in the conceptualization and design of this research. H-MC, I-CH, JP, JL, and SK were involved in data collection. H-MC, YEY, and G-YC analyzed the data, and J-BP, S-PL, Y-JK, and H-KK were responsible for investigation. SHS, HK, J-SL, and M-WS reviewed variant classifications. H-MC and S-HS wrote the manuscript draft and prepared figures. H-KK and M-WS reviewed and corrected the manuscript. All authors approved the final version of the manuscript.

## Funding

This research was funded by the Seoul National University Bundang Hospital, Republic of Korea [grant number: 06-2020-0130]. The funder had no role in study design, data collection and analysis, decision to publish, or preparation of the manuscript.

## Disclosures

HK Kim is a consultant for BMS and receives research grant from BMS and Chon Kun Dang.

## Acknowledgment

None

## Abbreviations

ACMG/AMP: American College of Medical Genetics and Genomics
CMR: Cardiac magnetic resonance
CV: Cardiovascular
HCM: Hypertrophic cardiomyopathy
ICD: Implantable cardioverter-defibrillator
LVOT: Left ventricular outflow tract
NGS: Next-generation sequencing
SCD: Sudden cardiac death
VT: Ventricular tachycardia
VUS: Variant of uncertain significance

## Notes

### Competing Interest Statement

The authors have declared no competing interest.

### Author Declarations

The study protocol was approved by the Institutional Review Board of Seoul National University Hospital (IRB No. J-2602-013-1713) and Seoul National University Bundang Hospital (IRB No. B-2004-604-409), with a waiver of written informed consent due to the retrospective study design.

